# Computational and Experimental Antibody Affinity and Diagnostic Accuracy Quantification of SARS-CoV-2 SD2 Major Disulfide Loop Analog

**DOI:** 10.64898/2026.06.05.26353587

**Authors:** Brian Andrich L. Pollo, Glenmarie Angelica Perias, Riziel Hannah Aguimatang, Ayra Patrice Espiritu, Danica Ching, Maria Isabel Idolor, Ruby Anne King, Fresthel Monica Climacosa, Salvador Eugenio Caoili

## Abstract

**Introduction:** Synthetic oligopeptides provide a rapid and cost-efficient approach to developing antibodies and diagnostics for emerging viral variants.

**Methods:** This study computationally and experimentally characterized a synthetic peptide analog of the SARS-CoV-2 spike subdomain 2 major disulfide loop (SD2MDL), designated S621 (CPVAIHADQLTPTWRVYSTC). Binding affinity was computationally estimated using the Heuristic Affinity Prediction Tool for Immune Complexes (HAPTIC), while experimental validation was performed using enzyme-linked immunosorbent assay (ELISA) with rabbit-derived antipeptide antibodies. Clinical diagnostic accuracy testing was done using plasma samples from RT-PCR– confirmed COVID-19 patients and pre-COVID-19 controls.

**Results:** S621 demonstrated nanomolar binding affinity 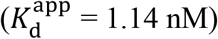 and high avidity (3.67 nM), closely matching HAPTIC predictions (3.54 nM). Diagnostic evaluation yielded a sensitivity of 89.92% and specificity of 27.79%, corresponding to an overall accuracy of 71.79%.

**Discussion:** These findings demonstrate that a single synthetic peptide derived from a conserved spike subdomain can function as a high-affinity surrogate for full-length antigens, supporting its potential application in rapid peptide-based immunodiagnostics.

## 1 Introduction

The global experience with COVID-19 has emphasized urgency in quickly developing and testing diagnostics and therapeutics (1). Since its identification in late 2019, SARS-CoV-2 has evolved into multiple variants, each with altered antigenic properties, thus affecting the performance of existing vaccines and complicating serological surveillance (2). Vaccines based on the spike protein have demonstrated substantial protection, yet the production of full-length recombinant proteins requires extensive infrastructure, purification steps, and temperature-controlled storage (3). These limit the speed at which new biotechnological products can be generated in response to viral evolution.

In response to this, synthetic oligopeptides serve as alternatives, combining specificity with chemical simplicity (4,5). By reproducing defined antigenic regions, peptides can provide targets for antibody detection or vaccine design without the logistical constraints of full-length proteins (6). Chemical synthesis allows rapid modification of sequences and the incorporation of stabilizing features (7). Multivalent constructs, achieved through polymerization or crosslinking, can simulate epitope clustering, enhancing apparent antibody binding through avidity (8). Despite these advantages, peptides have lower intrinsic binding affinity compared with native proteins (9). Linear sequences may fail to capture conformational epitopes, and the choice of sequence, chemical modifications, and presentation format all influence assay performance.

Research on SARS-CoV-2 has focused primarily on peptides derived from spike or nucleocapsid proteins (10,11). Conserved loops in subdomain 2 of the spike protein have been previously identified in structural studies as both accessible to antibodies and relatively conserved across variants (12). Computational methods can prioritize sequences likely to engage antibodies effectively, yet most studies do not combine *in silico* predictions with experimental validation and testing in clinical samples. The lack of an integrated workflow limits the translation of peptide candidates into practical diagnostic tools and hinders assessment of their performance in diverse patient populations.

This study aimed to determine whether synthetic peptides derived from structurally conserved regions of SARS-CoV-2 spike protein can serve as functional surrogates for full-length antigens in diagnostic applications. The focus is on the major disulfide loop of subdomain 2, a region predicted to be immunogenic and surface-exposed. Specifically, this study aimed to computationally and experimentally determine the affinity of the SARS-CoV-2 spike SD2 major disulfide loop (SD2MDL) toward cognate antibodies. Specifically the study aimed to develop immunogenic, conserved synthetic peptide analogs of SD2MDL, experimentally estimate the affinity of these analogs using antipeptide antibodies, and demonstrate the binding of said analogs to cognate antibodies in clinical samples.

## 2 Materials and Methods

### 2.1 Design and in silico evaluation of SD2MDL peptide analogs

The peptide analog **S621** (CPVAIHADQLTPTWRVYSTC) (**Table 1**) was designed by B-cell epitope prediction, with affinity computationally estimated using the Heuristic Affinity Prediction Tool for Immune Complexes (HAPTIC) (13). In brief, paratope-epitope binding affinity was expressed in terms of the standard free-energy change of binding 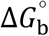:

**Table 1.**
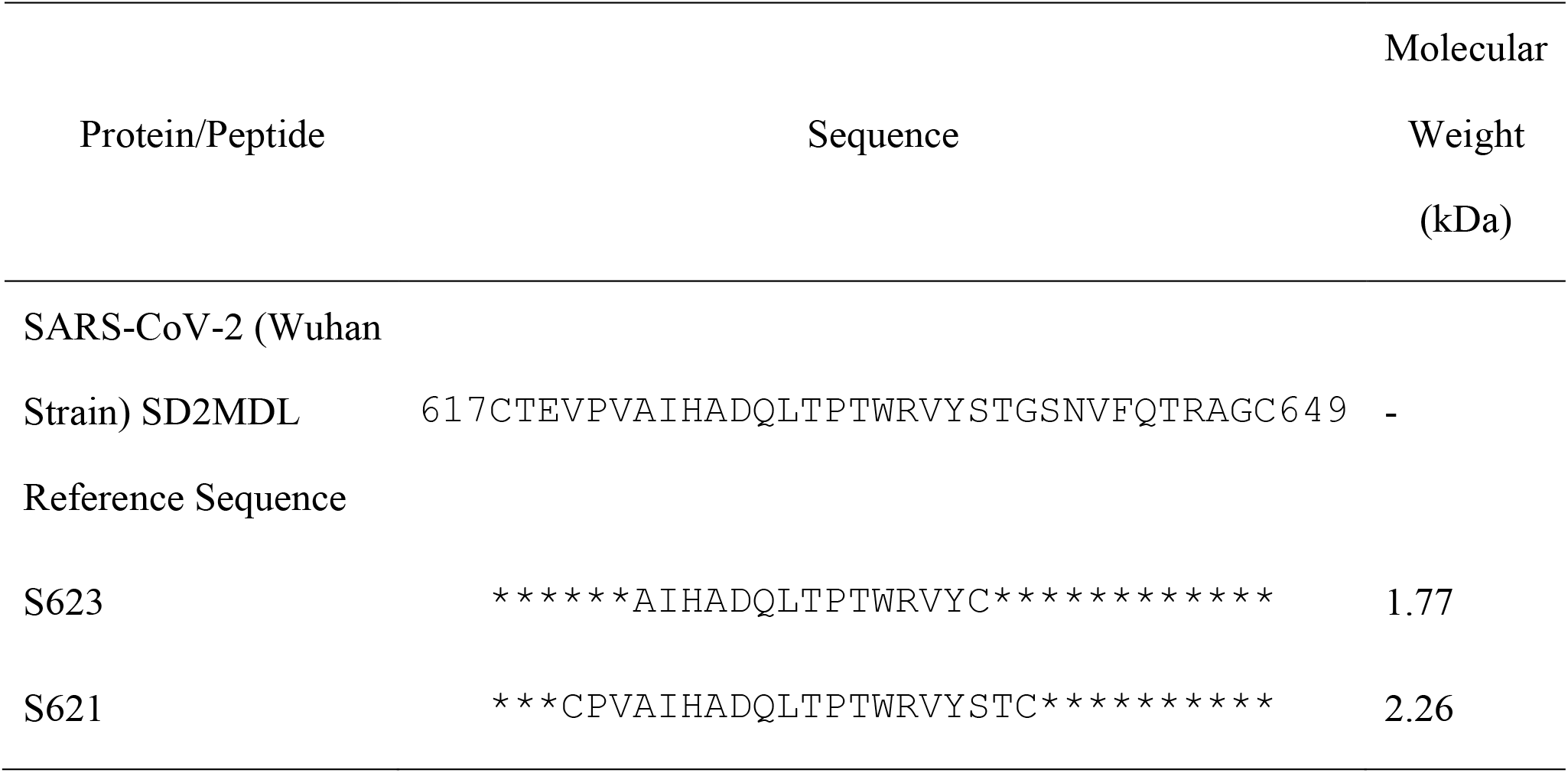
Sequences of Synthetic Peptide Antigenic Constructs (SPACs) representing SD2MDL and the SD2MDL region.

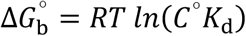

where *R* is the gas constant, *T* the absolute temperature, *C* the standard concentration (set by convention to 1M), and *K*_d_ the dissociation constant.

### 2.2 Peptide design

Structural and functional analyses were conducted by retrieving three-dimensional structures from the Protein Data Bank (PDB; https://www.rcsb.org/) and UniProt (https://www.uniprot.org/, RRID:SCR_002380), visualized using PyMOL (https://www.pymol.org/, RRID:SCR_000305). Sequence homology was assessed using BLASTp to ensure specificity to SARS-CoV-2 epitopes. Sequence-based mutation effects were analyzed using resources available through the ExPASy bioinformatics portal. Peptides were modified by adding cysteine residues at the N- and C-termini to facilitate polymerization through disulfide bond formation.

#### 2.2.1 Peptide synthesis and polymerization

The antigenic peptides were synthesized by Genscript (Singapore), a commercial provider. Solid-phase peptide synthesis was employed, and the peptides were analyzed for purity and identity using high-performance liquid chromatography (HPLC). The peptides were then subjected to polymerization under mild oxidizing conditions. Dimethyl sulfoxide (DMSO) was added to achieve a final concentration of 50%, and the mixture was incubated at room temperature for 24 hours to promote disulfide bond formation.

#### 2.2.2 Binding analysis of peptide–antibody interactions

To experimentally estimate *K*_d_, an in-house ELISA was performed on antipeptide antibodies (ApAbs) prepared by immunizing rabbits with **S623** (AIHADQLTPTWRVYC) and affinity purifying from the resulting sera. The resulting ELISA absorbance values were then plotted against antibody concentration and linearized according to the equation of Orosz & Ovádi (2002) (14):

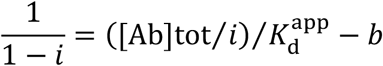

where *i* is the relative saturation, [Ab]tot the total antibody concentration, and 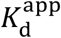 the apparent dissociation constant (calculated from the slope of the resulting linear plot).

#### 2.2.3 Generation and purification of anti-S621

New Zealand White rabbits were immunized with keyhole limpet hemocyanin (KLH)-conjugated S621 peptides emulsified in Freund’s adjuvant (Day 0, 14, 28). Sera were collected on Day 42 and affinity-purified on M1-Sepharose 4B columns.

#### 2.2.4 Peptide depolymerization

S621 was subjected to depolymerization to assess the contribution of the polymeric structure to avidity. N-acetylcysteine (Nac) was used as a reducing agent to break disulfide linkages. Aliquots of the polymeric peptide were incubated with various concentrations of Nac at 37°C for 30 minutes. To match the composition of the test solutions with the negative control, DMSO was added to a final concentration of 20%. The negative control was prepared by heating Nac in the presence of DMSO to degrade it. The complete degradation of Nac was confirmed using Ellman assay and by the evolution of the characteristic odor of dimethylsulfide. Peptide polymerization and depolymerization was confirmed with SDS-PAGE (**Supplementary figure 1**). The apparent dissociation constant 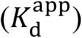 was calculated for both the polymeric and depolymerized forms of the peptide.

#### 2.2.5 S621 ELISA

To assess peptide-antibody interactions, indirect ELISA was performed as previously described in the detailed protocol available on protocols.io (15). Peptides were diluted in coating buffer to achieve a final concentration of 10 nM peptide and used to coat high-binding 96-well microtiter plates. The plates were incubated at 4°C overnight, followed by washing with phosphate-buffered saline containing 0.05% Tween-20 (PBST). Blocking was performed with 2% skimmed milk in PBST at 37°C for 30 minutes. Serial dilutions of anti-TNP antibody were then applied, followed by incubation with 1:2000 protein A peroxidase conjugate (0.5μg/mL). Chromogenic development was achieved using tetramethylbenzidine and hydrogen peroxide, and the reaction was halted with 0.1 M sulfuric acid. Absorbance was recorded at 450 nm, and 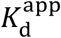 values were calculated from the slope of linearized plots.

### 2.3 Assessment of antibody recognition in clinical plasma

#### 2.3.1 Clinical samples

Clinical samples were prospectively collected from patients who satisfied the following eligibility criteria: (1) adults (≥18 years), (2) with RT-PCR-confirmed COVID-19, (3) admitted at Philippine General Hospital, the national referral hospital, between October 2020 and February 2021, prior to the national vaccine rollout. Discharged and absconded patients were excluded. Potential participants were identified by the attending physician using convenience sampling. Blood was drawn on days 1, 7, and 14 of hospitalization, coinciding with routine clinical sampling. All sera were heat-inactivated at 56 °C for 30 minutes and stored at −80 °C until use. Disease severity was classified based on Department of Health guidelines: asymptomatic (RT-PCR positive), mild (with symptoms but no pneumonia), moderate (with pneumonia), severe (requiring oxygen supplementation), or critical (requiring mechanical ventilation).

#### 2.3.2 Controls

Negative control sera were obtained from healthy blood donors prior to December 2019. Samples were treated identically to patient sera, including heat inactivation and storage conditions, to ensure assay consistency.

#### 2.3.3 Diagnostic accuracy determination

The result of the S621 ELISA was compared against the index test, RT-PCR as performed by non-blinded laboratory personnel, following standard operating procedures (cycle threshold: 40 or less). Clinical data was available to medical technologists and pathologists for interpretation. Diagnostic performance was evaluated using statistical software (MedCalc version 23.0.8, RRID:SCR_015044). The prespecified cutoff was determined by change-point analysis (16). The *post hoc* cutoff for ELISA positivity was determined via Youden index. Accuracy measures were calculated for both prespecified and exploratory thresholds. No indeterminate results were reported, and all samples were included in the analysis. Sample size estimates were determined using PASS software (version 20.0.7, RRID:SCR_019099) based on an expected prevalence of 42.3% and a binary outcome assumption (17,18). Figures were created using BioRender (https://BioRender.com, RRID:SCR_018361).

#### 2.3.4 Ethics

The study was conducted following the principles of the Declaration of Helsinki and the standards for reporting diagnostic accuracy (STARD) guidelines (19). Ethics approval was obtained from the University of the Philippines Manila Research Ethics Board (Code 2022-0492-01). The full protocol is registered under HERDIN (https://www.herdin.ph/) (PHRR230601-005772). All authors had access to study data and approved the final manuscript. Laboratory personnel performing ELISA were blinded to patient disease status throughout testing.

## 3 Results

### 3.1 Conservation and immunogenic features of SD2MDL analogs

The peptide analog S621 was selected to represent a conformationally flexible segment of the SD2 major disulfide loop (SD2MDL) of the SARS-CoV-2 spike protein (**Figure 1A**). Sequence alignment and structural mapping using PDB entries 6XR8 and 7CWL revealed that this segment is highly conserved across multiple variants, including Alpha, Beta, Gamma, Delta, Epsilon, and Omicron (**Figure 1B**). Notably, several residues within S621 were unresolved in the crystal structures, indicative of conformational disorder and surface exposure, features that are consistent with B-cell epitope accessibility. Computational B-cell epitope prediction using HAPTIC highlighted residues AIHADQLTPTWRVY as the most immunodominant within this segment. These analyses indicate that the selected peptide region preserves key structural and antigenic features across major viral lineages and is likely to retain antibody recognition potential despite variant-specific amino acid changes. Structural visualization highlighted the surface orientation of the loop and potential accessibility to circulating antibodies, with a notable absence of steric hindrance from neighboring S1 subdomains.

**Figure 1.**
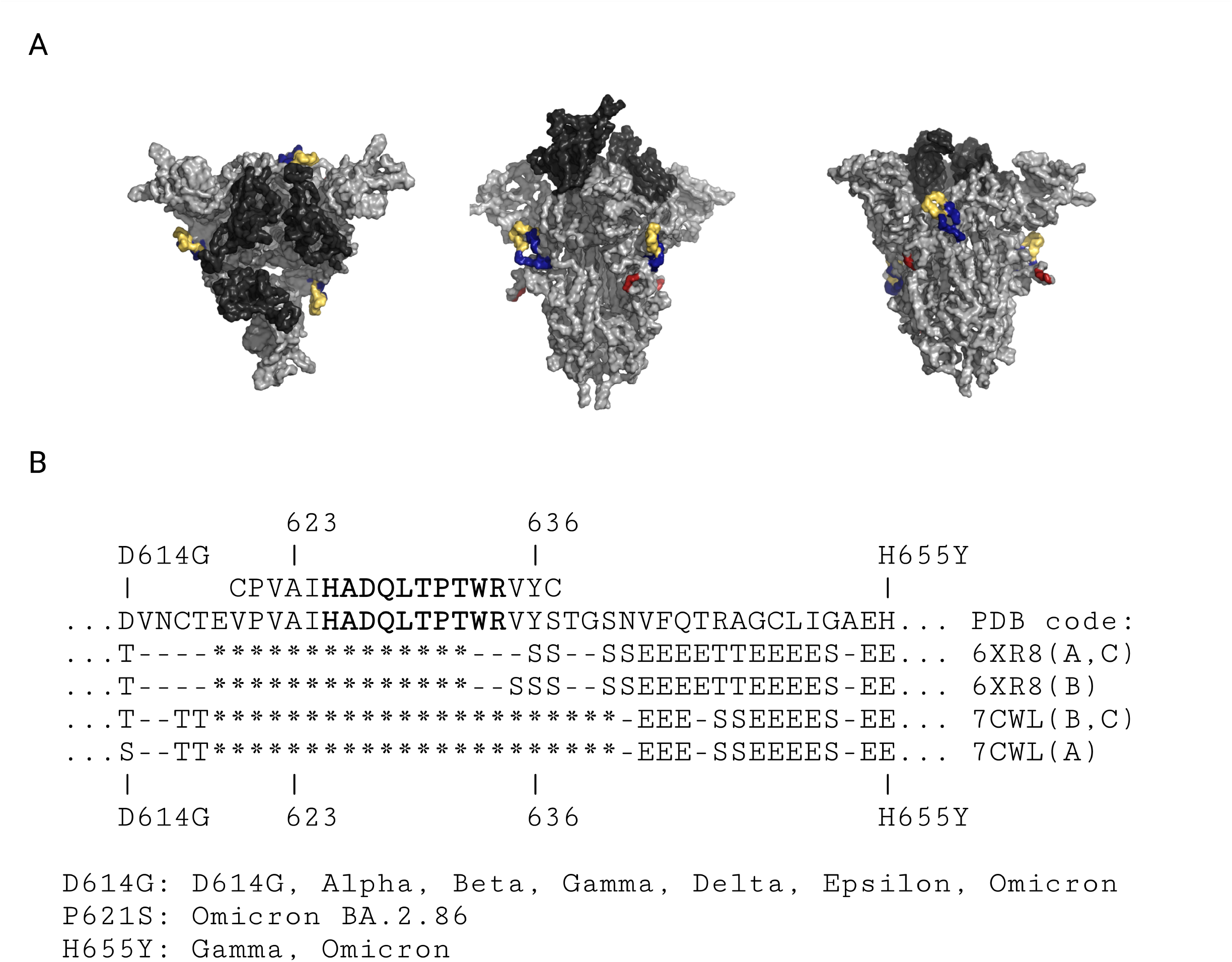
S621 was a peptide analog of an antibody-accessible, conformationally disordered sequence segment of SD2MDL that is conserved across various SARS-CoV-2 strains. (A) Three-dimensional structure of the SARS-CoV-2 spike glycoprotein (PDB ID: 7DZY). The S1 subunits are shown in yellow, the subdomain 2 major disulfide loop (SD2MDL) regions in blue, the receptor-binding domain (RBD) in black, and the S1/S2 cleavage sites in red. (B) Multiple S621 residues are unresolved by X-ray crystallography. Representative alignments of secondary-structure identifiers formatted as <PDB ID> (<chain>). Structural data were derived from X-ray crystallography (*: unassigned atomic coordinates, S: bend, E: extended strand, T: turn).

### 3.2 High-affinity interactions between peptide analogs and antibodies

The affinity of S621 for its cognate antibodies was examined both computationally and experimentally. HAPTIC-based *in silico* modeling predicted an apparent dissociation constant 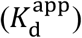 of 3.54 nM, reflecting strong interaction potential. Experimental evaluation using an in-house ELISA confirmed high-affinity binding, with a 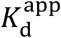 of 1.14 nM for monomeric interactions and an overall avidity of 3.67 nM for polymerized forms (**Figure 2**). The modest 3.54% deviation between predicted and measured values supports the accuracy of the computational affinity estimates. Observed binding curves demonstrated a typical sigmoidal pattern consistent with saturable, high-affinity interactions, with negligible background signal observed in negative control wells containing irrelevant peptides or pre-pandemic plasma samples. Comparative analysis between monomeric and polymerized S621 revealed a measurable avidity gain, suggesting that the multivalent presentation of epitopes enhances effective binding. he EC_50_ value of polymeric S621 was 5.048 (95%CI: 4.366 – 5.982), which was significantly lower than that of the monomeric/depolymerized form at 2.117 (95%CI: 1.985 – 2.263) nM, *p* < 0.0001, unpaired *t*-test), indicating a higher avidity. These findings provide quantitative evidence that polymerized S621 maintains structural integrity while promoting enhanced antibody recognition.

**Figure 2.**
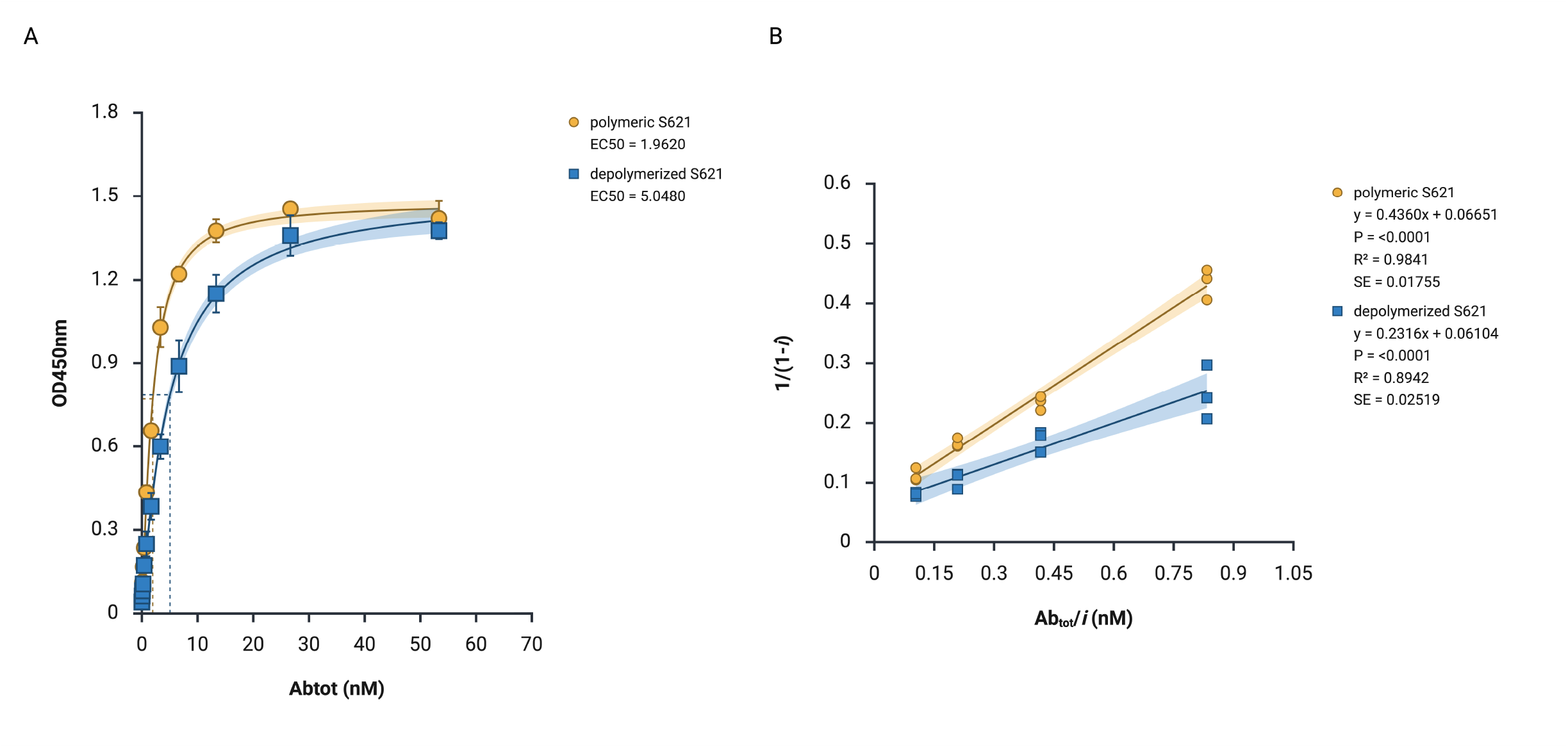
Computational and experimental *K*_d_ estimates were closely matched. Using in-house ELISA, binding affinity and avidity (*i*.*e*., overall affinity) was calculated to be 1.14 nM and 3.67 nM, respectively. HAPTIC estimated a binding affinity of 3.54 nM. This corresponds to a 3.54% error, suggesting that computational affinity estimation was accurate. (A) Indirect ELISA was used to measure immunocomplex formation of antipeptide antibodies with monomeric and with depolymerized form S621. (B) Plots of normalized ELISA absorbance readings against total antipeptide antibody concentration were used to estimate the apparent dissociation constant of the monomeric and the depolymerized form of S621.

### 3.3 Recognition of SD2MDL analogs by antibodies in clinical samples

The diagnostic performance of S621 was evaluated using plasma samples collected from 1,443 individuals with RT-PCR–confirmed SARS-CoV-2 infection prior to vaccination rollout (**Supplementary table 1, Figure 3A**). The ELISA incorporating S621 achieved a sensitivity of 89.92% (95% CI: 87.91–91.70%) and a specificity of 27.79% (95% CI: 23.56–32.34%) (**Figure 3B,C, Table 2**). Overall diagnostic accuracy was 71.79%, with positive and negative likelihood ratios of 1.25 and 0.36, respectively. Signal intensity was consistently higher in confirmed positive samples, whereas pre-pandemic negative controls exhibited low optical density readings, indicating minimal non-specific binding. Despite high sensitivity, the relatively low specificity suggests potential cross-reactivity with antibodies generated in response to related epitopes, warranting further evaluation with variant-specific panels. Subgroup analyses by age, sex, and disease severity showed no statistically significant differences in ELISA signal, supporting broad applicability across diverse patient populations.

**Table 2.** Clinical diagnostic performance of S621 ELISA.

| Statistic | Value | (95% CI) |
| --- | --- | --- |
| Sensitivity | 89.92% | (87.91% - 91.70%) |
| Specificity | 27.79% | (23.56% - 32.34%) |
| PLR | 1.25 | (1.17 - 1.33) |
| NLR | 0.36 | (0.29 - 0.46) |

**Figure 3.**
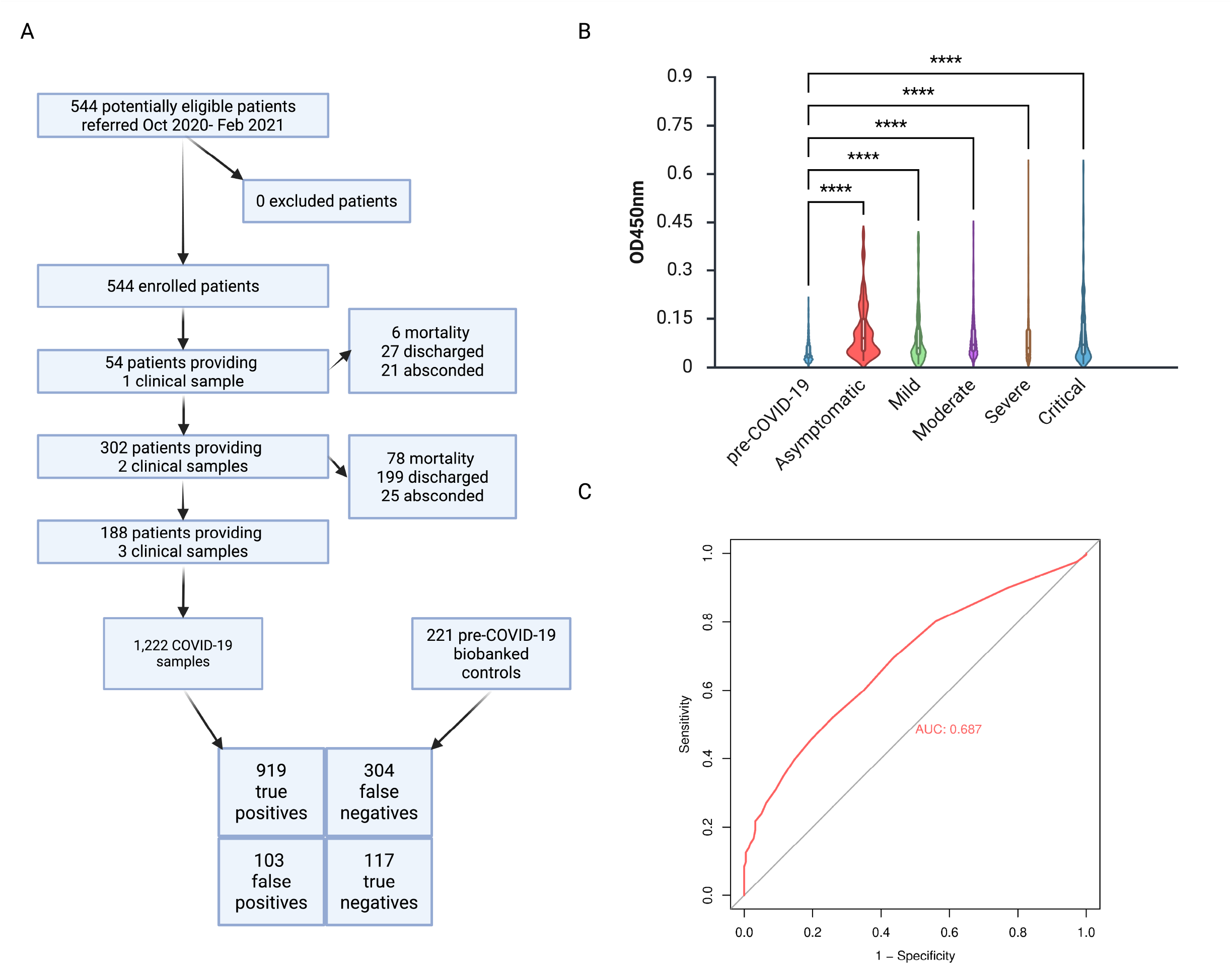
S621 binds clinical antibodies sensitively. S621 was subsequently developed into a diagnostic assay, and testing using predetermined thresholds on 1,443 samples and controls revealed a sensitivity of ~90%, corresponding to an accuracy of around 71.79%. (A) Patient flow diagram, (b) dot plot (one way ANOVA, Dunn’s multiple comparisons, ****: *p* < 0.0001), and (B) receiver operating characteristic (ROC) curve.

## 4 Discussion

The peptide analog S621 corresponds to a highly conserved and surface-exposed segment of the SD2 major disulfide loop of the SARS-CoV-2 spike protein. This region is preserved across major variants, with minimal substitutions affecting residues implicated in B-cell epitope recognition (**Figure 1B**), as demonstrated with sequence alignment and structural mapping. These findings are consistent with previous structural analyses suggesting that SD2 loops maintain a stable surface orientation across SARS-CoV-2 variants while exhibiting conformational flexibility (20). The unresolved residues in PDB entries 6XR8 and 7CWL correspond to regions likely to experience dynamic disorder, a characteristic that enhances antibody accessibility. This observation aligns with reports that conformationally disordered, flexible loops often serve as immunodominant epitopes due to their increased solvent exposure and adaptability in binding to diverse paratopes (21,22). The computational identification of residues AIHADQLTPTWRVY as a dominant B-cell epitope further validates the selection of S621 as a candidate diagnostic peptide. Collectively, these results emphasize the importance of targeting conserved, flexible regions when designing peptide-based diagnostics, as such epitopes retain cross-reactivity potential despite antigenic drift.

Experimental evaluation of S621 confirmed the high-affinity interactions predicted computationally. The apparent dissociation constant 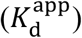 measured by ELISA closely matched the computationally derived estimate, demonstrating the reliability of *in silico* modeling using HAPTIC for pre-screening peptide candidates (**Figure 2**). The modest deviation between predicted and measured 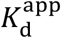 values may be attributable to differences between idealized computational models and the physicochemical environment *in vitro*, including multivalent interactions and potential peptide folding. Note than solvent effects are already taken into consideration in the calculation of HAPTIC. Polymerized S621 exhibited enhanced avidity relative to the monomeric form, indicating that multivalent presentation significantly improves effective binding. This finding is consistent with previous studies reporting that polymerization or clustering of epitopes can amplify apparent antibody affinity via avidity effects, a strategy successfully employed in both vaccine design and serological assays (23,24). The results suggest that computational pre-screening combined with polymerization strategies can accelerate the development of high-affinity peptide analogs suitable for diagnostic applications.

Application of S621 in clinical ELISA demonstrated strong sensitivity but limited specificity. The assay correctly identified 89.92% of confirmed positive samples, with negative controls yielding substantially lower signals (**Figure 3, Table 2**). The relatively low specificity (27.79%) is likely attributable to cross-reactivity with antibodies targeting structurally similar epitopes. This limitation highlights the trade-off between maximizing sensitivity and maintaining high specificity in peptide-based serological assays (25,26). These findings provide valuable context for the deployment of peptide-based diagnostics in surveillance and epidemiological studies, where sensitivity may be prioritized over specificity in early detection contexts.

The present findings highlight the feasibility of designing synthetic peptide analogs that recapitulate key immunogenic features of viral proteins. By targeting conserved, flexible loops, peptide-based diagnostics may retain effectiveness against emerging variants, addressing a critical limitation of conventional protein-based systems. Moreover, the demonstration that polymerization enhances avidity provides a methodological framework for optimizing peptide presentation in diagnostic assays. This approach may be extended to other viral antigens or conserved immunogenic regions in emerging pathogens, offering a scalable and modular alternative to recombinant protein-based diagnostics.

Several limitations must be acknowledged. The relatively low specificity observed for S621 necessitates further optimization, potentially through inclusion of multiple peptide epitopes. The study was performed on pre-vaccine sera, and performance may differ in post-vaccine populations or in samples with complex immunological histories. Quantitative comparisons of binding affinity relied on ELISA-derived K_d_^app^ measurements, which may not capture all kinetic parameters of peptide-antibody interactions. Future studies could incorporate surface plasmon resonance or biolayer interferometry for enhanced kinetic resolution Future studies should evaluate the performance of S621 in combination with complementary epitopes, assess longitudinal stability of antibody detection, and explore alternative multimerization strategies to further enhance specificity and avidity. Additionally, integration of peptide-based assays into high-throughput platforms may facilitate large-scale epidemiological surveillance and vaccine efficacy monitoring.

S621, a synthetic analog of the SARS-CoV-2 spike SD2 major disulfide loop, has nanomolar affinity toward cognate antibodies, shown both computationally and experimentally. Its strong binding and broad antibody recognition support its usefulness as surrogate for the full-length spike protein in immunodiagnostics. Work is underway to evaluate the binding of anti-S621 to intact SARS-CoV-2 virions. This oligopeptide-based strategy may be adapted for rapid development of locally produced immunodiagnostics, prophylactics, and therapeutic antibodies against endemic or emerging pathogens in the Philippines.

## 5 Conflict of Interest

The peptidic products (*i*.*e*., peptides and antipeptide antibodies) disclosed are covered by Philippine Patent no. 1/2021/050442 (WO 2023/022613, previously filed 13 August 2022), with coverage from 25 April 2025 to 13 August 2041. Letters patent for this invention was published on 5 June 2023.

## 6 Author Contributions

Writing – original draft: B.A.L.P.; writing – review and editing: B.A.L.P., G.A.S.P., R.H.B.A., A.P.N.E., D.S.C., M.I.C.I., R.A.N.K., F.M.M.C., S.E.C.C.; investigation: B.A.L.P., G.A.S.P, R.H.B.A., A.P.N.E., D.S.C., M.I.C.I., R.A.N.K.; conceptualization: B.A.L.P., F.M.M.C., S.E.C.C.; supervision: F.M.M.C., S.E.C.C..

## 7 Funding

This work was funded in part by the dissertation grant of B.A. L. P. from the Department of Science and Technology – Philippine Council for Health Research and Development (DOST-PCHRD).

## 8 Acknowledgments

Biobanked blood samples for testing was provided by the Section of Blood Banking and Transfusion Medicine, Department of Laboratories, University of the Philippines - Philippine General Hospital (UP-PGH). Sample collection was conducted in part alongside the “Collection and Archiving of Patient Sera and Plasma for Immunochemical Analysis to Detect Antibodies Against Infectious Pathogens (AbaCoV) Project” of DOST-PCHRD. Mr. Reneir John G. Tuason was involved in sample collection.

## 11 Supplementary Material

**Supplementary figure 1**. Polyacrylamide gel electrophoresis profile of S623 and S621 peptides.

**Supplementary table 1**. Baseline demographic and clinical characteristics.

## 12 Data Availability Statement

The datasets generated for this study can be found in the Harvard Dataverse, 2025, “Integrated In Silico, In Vitro, and Clinical Analysis of Antibody Binding to the SARS-CoV-2 Spike SD2 Disulfide Loop”, https://doi.org/10.7910/DVN/VHRNYF, Harvard Dataverse, V1

## Notes

**Conflict of interest statement: The authors declare a potential conflict of interest and state it below** The peptidic products (i.e., peptides and antipeptide antibodies) disclosed are covered by Philippine Patent no. 1/2021/050442 (WO 2023/022613, previously filed 13 August 2022), with coverage from 25 April 2025 to 13 August 2041. Letters patent for this invention was published on 5 June 2023.

### Author Declarations

Ethics approval was obtained from the University of the Philippines Manila Research Ethics Board (Code 2022-0492-01).

## References

1. Suri RK, Maugeais D, Maithal K. COVID-19 pandemic: A multidimensional analysis and the strategic role played by developing countries vaccine manufacturers. Vaccine (2025) 59:127271. doi: 10.1016/j.vaccine.2025.127271

2. Zabidi NZ, Liew HL, Farouk IA, Puniyamurti A, Yip AJW, Wijesinghe VN, Low ZY, Tang JW, Chow VTK, Lal SK. Evolution of SARS-CoV-2 Variants: Implications on Immune Escape, Vaccination, Therapeutic and Diagnostic Strategies. Viruses (2023) 15:944. doi: 10.3390/v15040944

3. Al-Fattah Yahaya AA, Khalid K, Lim HX, Poh CL. Development of Next Generation Vaccines against SARS-CoV-2 and Variants of Concern. Viruses (2023) 15:624. doi: 10.3390/v15030624

4. E. Sabaine A H. Castro-Kochi Ac, N. Mancini RS, A. Silva MR, F. Sepulveda A, R. Oliveira J, Remuzgo C, S. Santos K, L. Oliveira V, T. Kochi L, et al. Peptide-based biosensors for variant-specific detection of SARS-CoV-2 antibodies. Materials Advances (2025) 6:7090–7103. doi: 10.1039/D5MA00485C

5. Bulut A, Temur BZ, Kirimli CE, Gok O, Balcioglu BK, Ozturk HU, Uyar NY, Kanlidere Z, Kocagoz T, Can O. A Novel Peptide-Based Detection of SARS-CoV-2 Antibodies. Biomimetics (2023) 8:89. doi: 10.3390/biomimetics8010089

6. Li J, Ju Y, Jiang M, Li S, Yang X-Y. Epitope-Based Vaccines: The Next Generation of Promising Vaccines Against Bacterial Infection. Vaccines (Basel) (2025) 13:248. doi: 10.3390/vaccines13030248

7. Merrifield B. “3 - Solid-Phase Peptide Synthesis.,” In: Gutte B, editor. Peptides. San Diego: Academic Press (1995). p. 93–169 doi: 10.1016/B978-012310920-0/50004-8

8. Einav T, Yazdi S, Coey A, Bjorkman PJ, Phillips R. Harnessing Avidity: Quantifying the Entropic and Energetic Effects of Linker Length and Rigidity for Multivalent Binding of Antibodies to HIV-1. Cell Syst (2019) 9:466-474.e7. doi: 10.1016/j.cels.2019.09.007

9. Wang L, Wang N, Zhang W, Cheng X, Yan Z, Shao G, Wang X, Wang R, Fu C. Therapeutic peptides: current applications and future directions. Signal Transduct Target Ther (2022) 7:48. doi: 10.1038/s41392-022-00904-4

10. Yajima Y, Kosaka A, Ohkuri T, Hirohashi Y, Li D, Nagasaki T, Nagato T, Torigoe T, Kobayashi H. SARS-CoV-2 spike protein-derived immunogenic peptides that are promiscuously presented by several HLA-class II molecules and their potential for inducing acquired immunity. Heliyon (2023) 9:e20192. doi: 10.1016/j.heliyon.2023.e20192

11. Lee E, Sandgren K, Duette G, Stylianou VV, Khanna R, Eden J-S, Blyth E, Gottlieb D, Cunningham AL, Palmer S. Identification of SARS-CoV-2 Nucleocapsid and Spike T-Cell Epitopes for Assessing T-Cell Immunity. J Virol 95:e02002–20. doi: 10.1128/JVI.02002-20

12. Olukitibi TA, Ao Z, Warner B, Unat R, Kobasa D, Yao X. Significance of Conserved Regions in Coronavirus Spike Protein for Developing a Novel Vaccine against SARS-CoV-2 Infection. Vaccines (Basel) (2023) 11:545. doi: 10.3390/vaccines11030545

13. Caoili SEC. Prediction of Variable-Length B-Cell Epitopes for Antipeptide Paratopes Using the Program HAPTIC. Protein Pept Lett (2022) 29:328–339. doi: 10.2174/0929866529666220203101808

14. Orosz F, Ovádi J. A simple method for the determination of dissociation constants by displacement ELISA. J Immunol Methods (2002) 270:155–162. doi: 10.1016/s0022-1759(02)00295-8

15. Pollo BAL, King RA, Climacosa FMM, Caoili SEC. Synthetic peptide-based indirect ELISA using S1c (of the sequence CPVAIHADQLTPTWRVYSTC) as the antigen, per… (2025) https://www.protocols.io/view/synthetic-peptide-based-indirect-elisa-using-s1c-o-hb2eb2qbf [Accessed October 22, 2025]

16. Lardeux F, Torrico G, Aliaga C. Calculation of the ELISA’s cut-off based on the change-point analysis method for detection of Trypanosoma cruzi infection in Bolivian dogs in the absence of controls. Mem Inst Oswaldo Cruz (2016) 111:501–504. doi: 10.1590/0074-02760160119

17. Dispinseri S, Secchi M, Pirillo MF, Tolazzi M, Borghi M, Brigatti C, De Angelis ML, Baratella M, Bazzigaluppi E, Venturi G, et al. Neutralizing antibody responses to SARS-CoV-2 in symptomatic COVID-19 is persistent and critical for survival. Nat Commun (2021) 12:2670. doi: 10.1038/s41467-021-22958-8

18. Bujang MA, Adnan TH. Requirements for Minimum Sample Size for Sensitivity and Specificity Analysis. J Clin Diagn Res (2016) 10:YE01–YE06. doi: 10.7860/JCDR/2016/18129.8744

19. Cohen JF, Korevaar DA, Altman DG, Bruns DE, Gatsonis CA, Hooft L, Irwig L, Levine D, Reitsma JB, de Vet HCW, et al. STARD 2015 guidelines for reporting diagnostic accuracy studies: explanation and elaboration. BMJ Open (2016) 6:e012799. doi: 10.1136/bmjopen-2016-012799

20. Wong SWK, Liu Z. Conformational variability of loops in the SARS-CoV-2 spike protein. Proteins (2022) 90:691–703. doi: 10.1002/prot.26266

21. MacRaild CA, Richards JS, Anders RF, Norton RS. Antibody Recognition of Disordered Antigens. Structure (2016) 24:148–157. doi: 10.1016/j.str.2015.10.028

22. Novotný J, Handschumacher M, Haber E, Bruccoleri RE, Carlson WB, Fanning DW, Smith JA, Rose GD. Antigenic determinants in proteins coincide with surface regions accessible to large probes (antibody domains). Proceedings of the National Academy of Sciences (1986) 83:226–230. doi: 10.1073/pnas.83.2.226

23. Bennett NR, Jarvis CM, Alam MM, Zwick DB, Olson JM, Nguyen HV-T, Johnson JA, Cook ME, Kiessling LL. Modular polymer antigens to optimize immunity. Biomacromolecules (2019) 20:4370–4379. doi: 10.1021/acs.biomac.9b01049

24. O’Rourke JP, Peabody DS, Chackerian B. Affinity Selection of Epitope-based Vaccines using a Bacteriophage Virus-like Particle Platform. Curr Opin Virol (2015) 11:76–82. doi: 10.1016/j.coviro.2015.03.005

25. Klausberger M, Duerkop M, Haslacher H, Wozniak-Knopp G, Cserjan-Puschmann M, Perkmann T, Lingg N, Aguilar PP, Laurent E, De Vos J, et al. A comprehensive antigen production and characterisation study for easy-to-implement, specific and quantitative SARS-CoV-2 serotests. EBioMedicine (2021) 67:103348. doi: 10.1016/j.ebiom.2021.103348

26. Pagniez J, Petitdidier E, Parra-Zuleta O, Pissarra J, Bras-Gonçalves R. A systematic review of peptide-based serological tests for the diagnosis of leishmaniasis. Parasite 30:10. doi: 10.1051/parasite/2023011

