## Supplementary material for "Computational and Experimental Antibody Affinity and Diagnostic Accuracy Quantification of SARS-CoV-2 SD2 Major Disulfide Loop Analog"

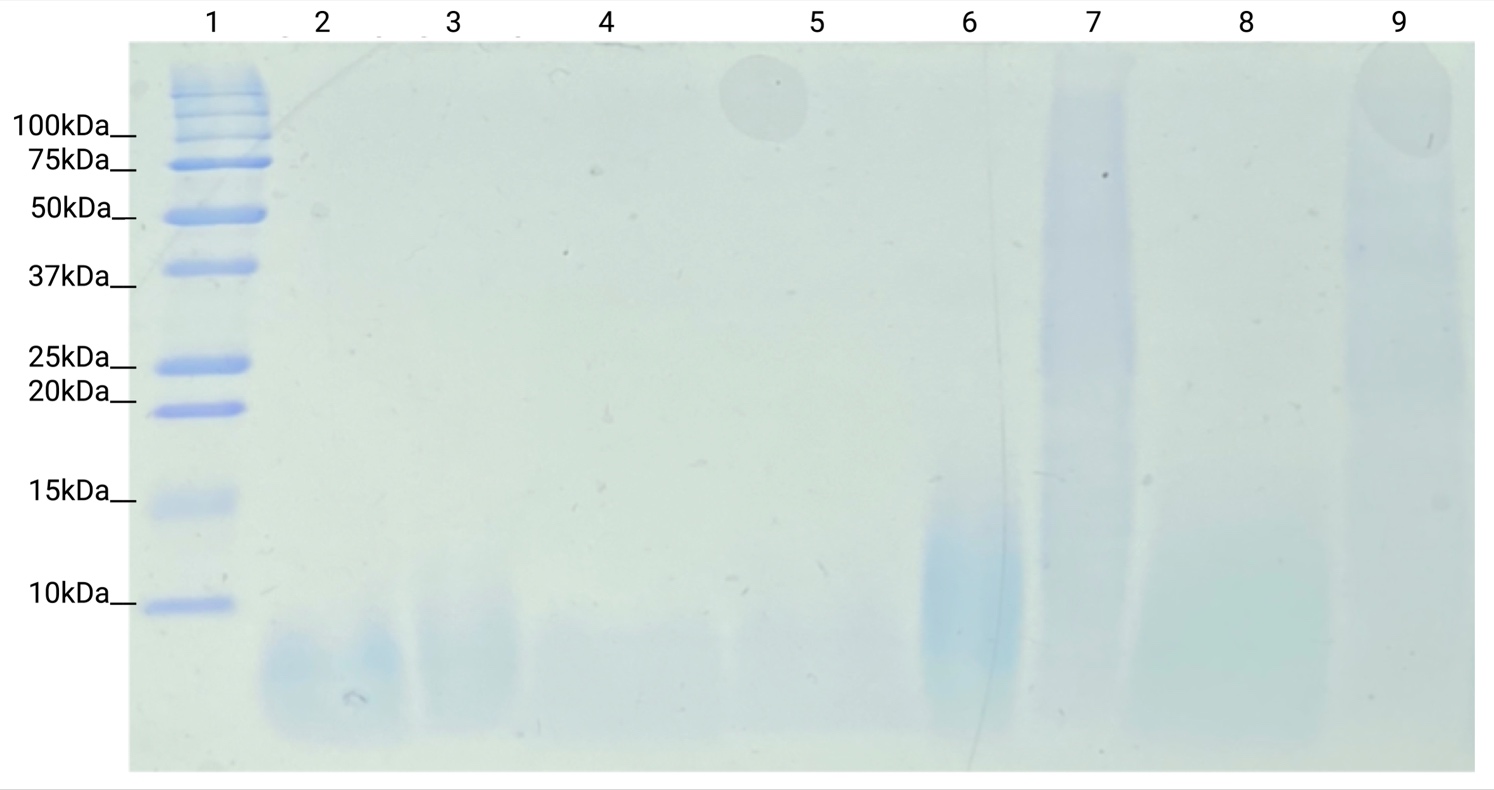

**Supplementary figure 1.** Polyacrylamide gel electrophoresis profile of S621 and S623. Peptides were oxidized in 50% DMSO for 1 or 24 hr, depolymerized using N-acetylcysteine. (1) MW ladder, (2,4) S623 depolymerized with equimolar N-acetylcysteine (in duplicate), (3,5) S621 depolymerized with equimolar N-acetylcysteine (in duplicate), (6) polymeric S623, (7) polymeric S621, (8) S623 depolymerized with 0.5:1 molar ratio N-acetylcysteine to peptide, (9) S623 depolymerized with 0.5:1 molar ratio N-acetylcysteine to peptide.

**Supplementary table 1.** Baseline demographic and clinical characteristics.

| **Characteristic** | **Value** |
| --- | --- |
| **Sex**^*^ |  |
| Male | 334 (61) |
| Female | 210 (39) |
| **Age** | ^†^54 ± 17 |
|  | ^‡^71 |
| **Disease severity**^*^ |  |
| Asymptomatic | 45 (8) |
| Mild | 47 (9) |
| Moderate | 119 (22) |
| Severe | 242 (44) |
| Critical | 91 (17) |

^*^ no. (%)
^†^ mean ± SD (years)

^‡^ range (years)
